# A randomized controlled trial of a video intervention shows evidence of increasing COVID-19 vaccination intention

**DOI:** 10.1101/2021.03.26.21254433

**Authors:** Leah S. Witus, Erik Larson

## Abstract

Increasing acceptance of COVID-19 vaccines is imperative for public health, as unvaccinated individuals may impede the ability to reach herd immunity. Previous research on educational interventions to overcome vaccine hesitancy have shown mixed effects in increasing vaccination intention, although much of this work has focused on parental attitudes toward childhood vaccination. In this study, we conducted a randomized controlled trial to investigate whether vaccination intention changes after viewing an animated YouTube video explaining how COVID-19 mRNA vaccines work. We exposed participants to one of four interventions – watching the video with a male narrator, watching the same video with a female narrator, reading the text of the transcript of the video, or receiving no information (control group). We found that participants who watched the version of the video with a male narrator expressed statistically significant increased vaccination intention compared to the control group. The video with a female narrator had more variation in results. As a whole, there was a non-significant increased vaccination intention when analyzing all participants who saw the video with a female narrator; however, for politically conservative participants there was decreased vaccination intention for this intervention, particularly at a threshold between being currently undecided and expressing probable interest. These results are encouraging for the ability of interventions as simple as YouTube videos to increase vaccination propensity, although the inconsistent response to the video with a female narrator demonstrates the potential for bias to affect how certain groups respond to different messengers.

**Significance Statement:** Widespread vaccination is important for ending the COVID-19 pandemic. This study investigates whether communicating the science behind new COVID-19 vaccines can increase people’s willingness to get vaccinated. We examined the effectiveness of an eight-minute animated video explaining how COVID-19 mRNA vaccines work, varying between a male narrator, a female narrator, and a control group. Participants who saw the video with a male narrator expressed a greater intent to get vaccinated than the control group. Participants who saw the video with a female narrator had more varied responses, including a decreased intent to get vaccinated among political conservatives. These findings indicate that science education may help increase vaccine uptake, but that beliefs about gender may influence how people receive such information.

## Introduction

Even before COVID-19 became a pandemic in 2020, vaccine hesitancy was considered a top global health threat by the World Health Organization (1). At the time this study was performed, in February 2021, Emergency Use Authorization had been issued by the United States Food and Drug Administration for two mRNA vaccines (Pfizer-BioNTech and Moderna) for the prevention of COVID-19, representing a tremendous public health success in their high efficacy and in the speed of their development. While vaccine production and access have been rapidly increasing in the United States and worldwide throughout the spring of 2021, overcoming vaccine hesitancy to achieve high rates of vaccine uptake may be important for achieving herd immunity (2, 3). Although studies have found that COVID-19 vaccine hesitancy has declined during the vaccine roll out (4), rates of vaccine hesitancy still remain concerningly high, particularly when unvaccinated individuals may pose a risk to members of society who are unable to be vaccinated (5). Therefore, research to collect empirical evidence on the effect of interventions to increase vaccination intention is needed.

This study examines an eight-minute, animated, educational YouTube video on the COVID-19 mRNA vaccines that we made called “COVID mRNA vaccines explained.” The video explains how the mRNA vaccine platforms work, highlights some of the positive features of the COVID-19 mRNA vaccines such as high efficacy and rarity of serious side effects, and emphasizes the altruism of vaccination. While the lack of editorial oversight of health information on YouTube and other social media platforms has contributed to the spread of vaccine misinformation and disinformation (6,7,8), YouTube videos could constitute an attractive tool for public health campaigns as they are easily disseminated through existing websites and social media platforms (9). Given the novelty of the COVID-19 vaccine platforms, and the urge to help end the pandemic through promoting vaccination, there are many health professionals, professors, and others who have been making and posting vaccine explainer videos for YouTube and other social media platforms such as Tiktok. However, there is little evidence about whether creating and sharing such videos is worthwhile: do these efforts actually affect vaccination propensity? Previous research on web-based educational interventions to increase vaccine acceptance, focused largely on parental attitudes toward childhood vaccinations, has found mixed results (10-12) – some studies found evidence of increased intention to vaccinate (13,14), some found no effect or effects that were not statistically significant (15,16), and some even reported a backfire effect wherein the intervention decreased vaccination intention (17,18). The prior literature highlights the need to continue such studies to identify effective interventions for overcoming vaccine hesitancy (11,19-21).

Herein we present the results of a randomized controlled trial conducted with 1,184 Mechanical Turk participants to investigate the effect on vaccination intention of an animated YouTube video explaining how the COVID-19 mRNA vaccines work. We investigated whether the exact same information presented in different formats would alter the efficacy of the communication by exposing treatment groups to either: a) watching the video with a male narrator (*Male-Narr*-Video https://youtu.be/Fv5bs4SPiYE), b) watching the same video with a female narrator (*Female-Narr*-Video https://youtu.be/j3hTeDyvgPs), c) reading a blog post containing the text of the transcript of the video, or d) receiving no information to serve as a control group. The visuals and scripts for both versions of the videos were identical; only the voice of the narrator differed. The results showed statistically significant higher vaccination intention in the group that watched the *Male-Narr*-Video compared to the control group, a robust association for a variety of alternative specifications. However, despite participants rating the quality of instruction of both the *Male-Narr*-Video and *Female-Narr*-Video equally highly, we found less consistent associations on intention to vaccinate for participants exposed to the *Female-Narr*-Video intervention. As a whole, the *Female-Narr*-Video produced a non-significant increase in vaccination intention compared to the control group. However, the impact on vaccination intention of the *Female-Narr*-Video varied by political identity of the participants. Political conservatives who were exposed to the *Female-Narr*-Video were less likely to express probable or definite interest in getting vaccinated than both other respondents exposed to the *Female-Narr-*Video and conservatives exposed to no information in the control group. Adjusting for this conditional association showed that for respondents who did not identify as politically conservative, both videos associated with increased vaccine intention at much closer rates, no matter the narrator. No such conditioning among political conservatives was observed for participants exposed to the *Male-Narr*-Video. Although further research is warranted to investigate the variation in efficacy based on the gender of the narrator, overall, this evidence supports the idea that educational YouTube videos, such as the *Male-Narr*-Video, may serve as an easy to share, simple way to increase vaccination intention.

### Video development

The YouTube video was made by LSW in December 2020, shortly after the issuance of Emergency Use Authorizations by the United States Food and Drug Association for the Pfizer-BioNTech and Moderna vaccines for COVID-19, which are both mRNA vaccines. The programs Biorender and Vyond were used to create the video, which uses animations and cartoon characters to illustrate educational points about how the COVID-19 mRNA vaccines work. The video begins with a review of the central dogma of biology, then explains how weakened virus vaccines work before explaining what an mRNA vaccine means. The video then emphasizes that the Pfizer-BioNTech (referred to as Pfizer in the video) and Moderna vaccines have been tested for safety and efficacy, and ends with an explanation of how getting vaccinated can protect others in society. Previous research on vaccine hesitancy interventions has identified common features for successful communication. This video incorporates many of these features, including: *i)* The video emphasizes the messages that approved vaccines are safe and go through evaluations (9), and that immunization also protects others, which has been found to activate positive emotions (2). *ii)* The video does not focus on messages that have been reported to impede efforts to diminish vaccine hesitancy such as myth-busting, fear-based, or numerically focused messages (10). *iii)* The video creates clear bottom-line (gist) messages through explanations of concepts in simple language rather than scientific jargon, the use of animated illustrations to visually reinforce the concepts explained verbally, and the use of text on the screen to underscore take-home messages. The Fuzzy-Trace theory of medical decision making has found that gist messages contribute to decision-making in a meaningful way relative to statistics and verbatim information (6, 7,10) and previous research on vaccine communication has found that articles with gist were shared on social media more than those without (22).

Some characteristics of the video are not as directly tied to recommendations from previous literature on vaccine interventions, partly because there is not yet much evidence on interventions specific for the COVID-19 vaccines. In some respects, the unique characteristics of the COVID-19 vaccine may alleviate some of the attitudes that have contributed to parental vaccine hesitancy for childhood vaccinations. For instance, it has been hypothesized that the benefits of some vaccinations may not seem tangible due to unfamiliarity with the harm of the preventable disease (9,23), which may be less likely for COVID-19. However, there are new concerns particular to COVID-19 vaccines that have been reported (2,21), and the video does address some of these, including unease over the new vaccine platforms that have not been used before, and concern over the speed of the vaccine development and approval (the video does not go into all of the details that contributed to the rapid vaccine development but does explain that the mRNA platform may allow quicker vaccine development than weakened virus platforms). Although previous research has shown that vaccine hesitancy is often more complex than a simple knowledge deficit (10,19), the overall tone of the video is an educational approach to influencing vaccination attitudes because in the case of the COVID-19 vaccines, the lack of knowledge about the new vaccine platform contribute to hesitancy.

Previous research has indicated that the messaging matters in addition to the message (24,25), and has recommended that vaccine interventions use an enthusiastic tone (10) and establish the public health educator as an expert (7). In this video, background music is used to create an engaging and enthusiastic tone. Although the video does not specify the credentials of the creator, the survey participants were told that the video was created by a college biochemistry professor. This study tests the effectiveness of a video that draws upon the best evidence in the vaccine hesitancy intervention literature for the message (information content) and messaging (style of communication) of vaccine communications, and additionally allows us to study whether there may be different patterns of effectiveness based on the messenger (the person who delivers the message). Since the narrator is never depicted in the video, we saw the video as a platform to isolate and test the effects of a single variable related to the messenger: the gender of the narrator’s voice. Therefore, two versions were used in the study, identical but for the voice of the narrator: the *Female-Narr*-Video version had a female voice reading the transcript and the *Male-Narr*-Video version had a male voice. In both cases the timing of the narration was matched to the animations (which led to a 3 second difference in length of the videos). The *Female-Narr*-Video was the original version created in December 2020 for educational purposes (although the title slide and link on YouTube were changed for this study), and the *Male-Narr*-Video was created later as an additional experimental condition. In other words, the creation of the *Female-Narr-*Video was independent of the idea of testing whether narrator gender might associate with its effectiveness; only after receiving positive anecdotal feedback on the original video did we decide to study the efficacy of such variations.

### Survey design

Participants completed a survey to measure COVID-19 vaccination intention, attitudes, and understanding. Many of the survey items were similar to validated items from the literature, with some adaptations since many of the previous scales of vaccine hesitancy have been used to measure parental attitudes towards childhood vaccinations (26). Using the Qualtrics survey platform, participants were randomly equally distributed into one of 4 paths: In paths 1, 2, and 3 the consent information page was followed by a page where participants watched the *Male-Narr*-Video, the *Female-Narr*-Video, or read the text of the video transcript presented as a blog post. On each of these pages, the time the participants spent on the page was recorded and advancing to the next page was disabled for a few minutes (approximately for half of the length of the video and two minutes for the approximately 1,200 word text) to increase the likelihood that participants engaged the material. After the information was presented in the video or text format, the participants encountered a section with questions that first asked about their intent to get vaccinated and then asked about their attitudes concerning the COVID-19 vaccines. The participants in the 4^th^ path did not receive any information and went directly to the vaccine intention and attitudes section. The vaccination intention item was based on the Imperial College London “Global attitudes towards a COVID-19 vaccine” survey (4) and asked “If a COVID-19 vaccine were made available to you this week, would you get it?” Response categories were: (1) Definitely no; (2) Probably no; (3) Undecided as of now; (4) Probably yes; (5) Definitely yes; and (6) I already got one or more dose of a COVID19 vaccine. This section also included questions on multiple domains related to vaccine hesitancy attitudes: a) perceptions of vaccine safety and efficacy, b) trust in medical, scientific, governmental and pharmaceutical authorities, c) common vaccine misconceptions and d) understanding of elements of vaccination (see SI for full survey questions)^†^. The next section of the survey asked about characteristics of the video or blog post, which included an attention check question, questions about the enjoyment of the video/blog post, and how likely participants would be to share it on social media, and was only shown to paths 1-3. This section also asked participants to rate the quality of instruction by the narrator and on the narrator’s trustworthiness, comfort and knowledge. All paths of the survey then went to a block of demographic questions and ended with a code to enter for payment on Amazon Mechanical Turk.

### Experimental set up

The survey was administered to 1,632 participants on Amazon Mechanical Turk on February 25^th^ and 26^th^, 2021 when the two approved COVID-19 vaccines in the United States were the Pfizer-BioNTech and Moderna mRNA vaccines. The only worker qualification required was that the Turk worker be located in the United States. Mechanical Turk is well-validated and considered a reliable source for survey data (27-29). The pay was $5, based on a $15/h wage and a 20-minute estimate of the average completion time. The study was approved by the Macalester College Institutional Review Board (Approval #022103). Participants were provided with consent information at the beginning of the study, although signed consent forms were not collected to preserve the anonymity of the participants. The dataset used for this analysis is available in the SI Appendix. Unless otherwise noted, the exclusion criteria that were applied were: exclusion of participants who had indicated they had already been vaccinated, exclusion of participants who answered the attention check question incorrectly, and exclusion of participants who spent less than 7 min on the video page (for a 7 min 37 sec video) or who spent less than 2 min 5 sec on the blog post page (which excluded a similar percentage of people for spending insufficient time as were excluded for insufficient time for the video exposures). These exclusion criteria resulted in n = 270 for the *Male-Narr*-Video group, n = 255 for the *Female-Narr*-Video group, n = 282 for the text group, and n = 393 for the control group.

## Results and discussion

### Data treatment

Since vaccination attitudes exist on a continuum with anti-vaccine refusal on one end, active demand on the other, and uncertainty and hesitation in the middle (30,31), we considered vaccine intention as a continuum rather than a simple hesitant/intent binary. Additionally, following previous research, we considered the possibility that the effects of an intervention may differ at distinct parts of the continuum (31). This presumption of asymmetric effects is also consistent with the idea that vaccine intent could be conceived as a series of stages, separated by thresholds, as noted in health research (32). Given this understanding, we modeled how exposure to each intervention associated with vaccine intention by using partial proportional odds models with the *gologit2* program in Stata using the program’s autofit option (33,34). These models preserve the order across the categories of vaccine intent while also allowing for either a single coefficient for a variable across the different levels of intent or for separate coefficients for each level of intent. Thus, some variables may associate with a consistent change in moving from one level to the next level of vaccine intention across different levels of the continuum, while other variables may have asymmetric associations, such as varying levels of intensity or direction at different levels of vaccine intention. The findings discussed below were robust to a variety of alternative specifications, as described in the SI Appendix.

Although participants were randomly assigned to treatment conditions, we included a series of control variables (respondents’ race/ethnicity, gender, political ideology, age, and education) in our models that may associate with vaccine intention. Mechanical Turk respondents are a diverse, but not nationally representative group (28) (see Supplemental Table 1 for the frequency of these variables). We excluded variables for which there were no significant associations in the multivariate models for the sake of parsimony. We first estimated a model with the treatment variables and control variables (Model 1), and then estimated a model testing whether the association between *Female-Narr*-Video and vaccine intent varied based on respondents identifying as conservative (Model 2). Table 1 shows the results from these two models. In both models, the race/ethnicity variables are Black and Native American compared to all others who identify as neither. While some studies suggest that Hispanic/Latinx individuals may exhibit lower vaccine intent (35,36), and while Black, Indigenous, and Hispanic/Latinx populations have all experienced histories of medical abuse that may decrease current medical trust (37,38), we found no association between Hispanic/Latinx and vaccine intent in our analysis.^‡^ In addition, we included separate variables for people who described themselves as politically conservative and people who described themselves as politically liberal, comparing them to those identified politically as neither conservative nor liberal (political moderates and those who identify as something else). While trust in science is politicized, studies find differences in how political identities relate to such trust (41,42). Although political liberals may generally trust science more than others (42), conservatives may express greater distrust than others, linked more to cultural division and collective identity (43,44), suggesting that both groups should be examined in relation to moderates and others.

**Table 1.** Partial proportional odds ordered logit models for intention to vaccinate.

|  | Model 1 |  |  | Model 2 |  |  |
| --- | --- | --- | --- | --- | --- | --- |
|  | Coef. | s.e. | p | Coef. | s.e. | p |
| *Male-Narr-Video intervention* | 0.440 | 0.158 | .005 | 0.440 | 0.157 | .005 |
| *Female-Narr-Video intervention* | 0.312<sup>a</sup><br>0.069<sup>b</sup><br>-0.144<sup>c</sup><br>0.304<sup>d</sup> | 0.278<br>0.209<br>0.182<br>0.170 | .261<br>.741<br>.429<br>.073 | 0.374 | 0.191 | .051 |
| *Female-Narr-Video intervention x Conservative* |  |  |  | -0.444<sup>a</sup><br>-0.508<sup>b</sup><br>-0.971<sup>c</sup><br>-0.278<sup>d</sup> | 0.390<br>0.335<br>0.315<br>0.319 | .255<br>.129<br>.002<br>.383 |
| *Text intervention* | 0.168 | 0.155 | .278 | 0.175 | 0.154 | .255 |
| *Education: B.A. or Higher* | 0.911<sup>a</sup><br>1.088<sup>b</sup><br>1.160<sup>c</sup><br>0.643<sup>d</sup> | 0.209<br>0.164<br>0.146<br>0.134 | <.001<br><.001<br><.001<br><.001 | 0.909<sup>a</sup><br>1.100<sup>b</sup><br>1.186<sup>c</sup><br>0.638<sup>d</sup> | 0.210<br>0.163<br>0.146<br>0.134 | <.001<br><.001<br><.001<br><.001 |
| *Black* | -0.452 | 0.185 | .015 | -0.465 | 0.186 | .012 |
| *Native American* | -0.986 | 0.428 | .021 | -1.017 | 0.428 | .017 |
| *55 years or older* | 0.022<sup>a</sup><br>0.278<sup>b</sup><br>0.100<sup>c</sup><br>0.601<sup>d</sup> | 0.294<br>0.242<br>0.205<br>0.190 | .940<br>.250<br>.627<br>.002 | 0.083<sup>a</sup><br>0.334<sup>b</sup><br>0.148<sup>c</sup><br>0.574<sup>d</sup> | 0.294<br>0.240<br>0.203<br>0.189 | .777<br>.163<br>.468<br>.002 |
| *Female respondent* | -0.326 | 0.116 | .005 | -0.323 | 0.116 | .005 |
| *Liberal* | 1.251 | 0.151 | <.001 | 1.260 | 0.152 | <.001 |
| *Conservative* | -0.225 | 0.154 | .143 | -0.119 | 0.165 | .472 |
| *Constant* | 1.472<sup>a</sup><br>0.588<sup>b</sup><br>0.056<sup>c</sup><br>-0.943<sup>d</sup> | 0.216<br>0.191<br>0.185<br>0.186 | <.001<br>.002<br>.764<br><.001 | 1.438<sup>a</sup><br>0.506<sup>b</sup><br>-0.037<sup>c</sup><br>-0.969<sup>d</sup> | 0.214<br>0.191<br>0.185<br>0.187 | <.001<br>.008<br>.841<br><.001 |
|  | Wald chi-square 226.65<br>df 19 |  |  | Wald chi-square 234.36<br>df 20 |  |  |

N=1,184
Dependent variable coding: (1) Definitely no; (2) Probably no; (3) Undecided as of now; (4) Probably yes; (5) Definitely yes.
For variables that meet the proportional odds assumption, there is one coefficient for all comparisons between levels of vaccination intention
For variables that violate the proportional odds assumption and constant:
<sup>a</sup>Coefficient for any response with higher intention to vaccinate than “definitely no” response;
<sup>b</sup>Coefficient for any response with higher intention to vaccinate than “probably no” response;
<sup>c</sup>Coefficient for either yes response compared to no or undecided responses;
<sup>d</sup>Coefficient for “definitely yes” response compared to any lower intention to vaccinate responses.

### Findings

Model 1 shows a positive association between having been exposed to the *Male-Narr*-Video and vaccination intent, consistent across all levels of intent. Compared to the control group given no information, respondents who watched the *Male-Narr*-Video were more likely to express vaccination intention as measured by their response to the question how likely they would be to receive a COVID-19 vaccine if it were available to them this week (coefficient = 0.440, *p* = 0.005). Figure 1 illustrates the substantive meaning of these coefficients by displaying predicted probabilities for a respondent who is white, male, politically moderate, under 55, and with at least a B.A. degree in each of the four experimental conditions (45). Such a respondent exposed to the *Male-Narr-*Video would have a predicted probability of 53.5% of replying “definitely yes” for their intention to be vaccinated, compared to a predicted probability of 42.6% for the control group. Those exposed to the text intervention had a much smaller, statistically insignificant increase in vaccination intention compared to the control group (coefficient = 0.168, *p* = 0.278). The pattern for respondents exposed to the *Female-Narr*-Video intervention, however, had a more nuanced pattern, as there were asymmetric associations across the different levels of vaccine intent. Although outside of the standard cut-off for statistical significance, those exposed to the *Female-Narr*-Video had a greater likelihood of answering “definitely yes” to getting the vaccine compared to the control group (coefficient = 0.304, *p* = 0.073), albeit to a lesser degree than those exposed the *Male-Narr*-Video. Again, although not statistically significant, the negative coefficient for expressing a response with higher intention to vaccinate than “undecided” among those exposed to the *Female-Narr*-Video is striking (coefficient = –0.144, *p* = 0.429). This pattern suggests that there may have been heterogeneous effects of being exposed to the female video, which we tested in Model 2.

**Figure 1.**
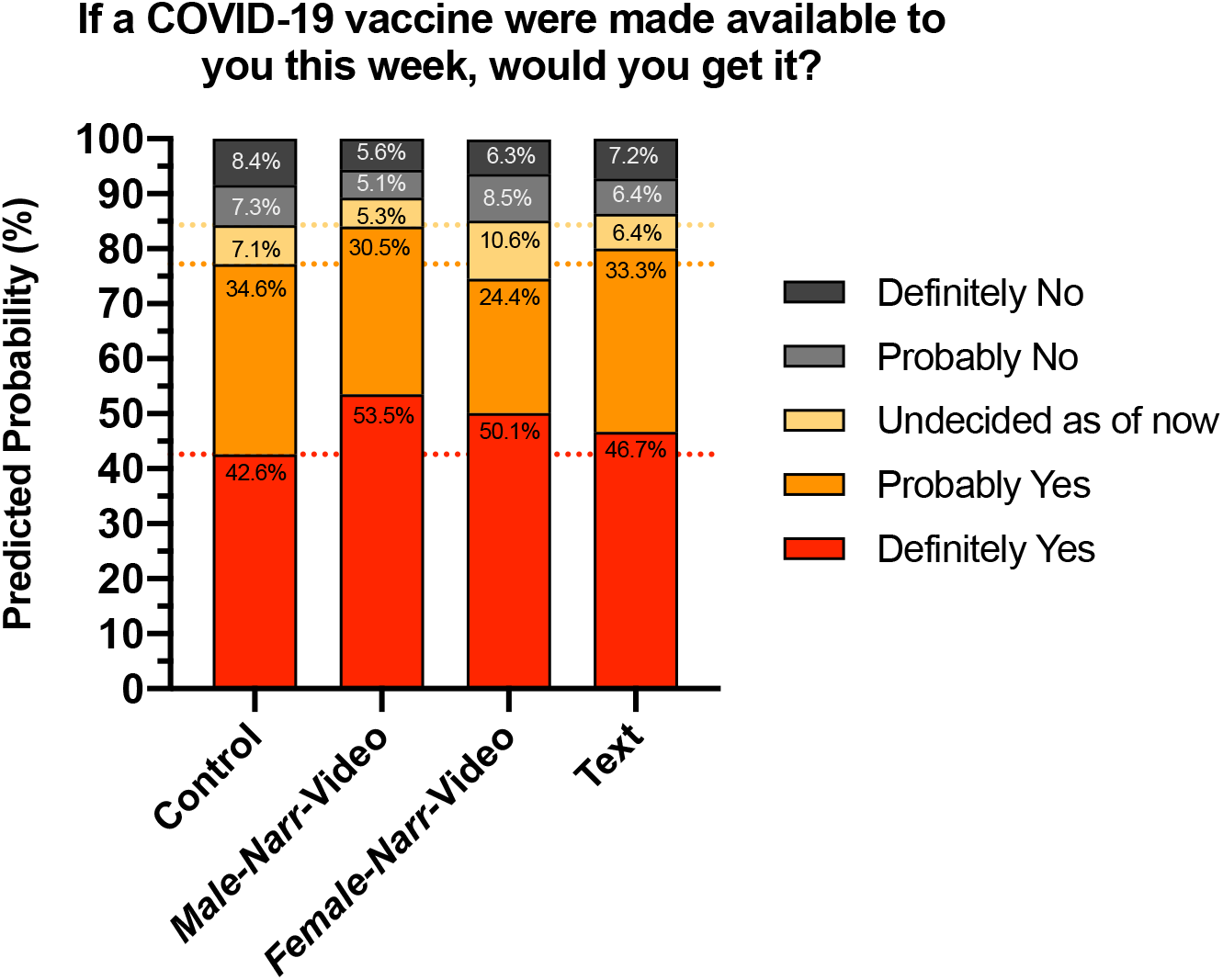
Predicted probabilities for a participant who is white, male, political moderate, under 55, and with at least a B.A. degree upon exposure to each of the four interventions, illustrating the substantive meaning of the coefficients of Model 1. The horizontal dotted lines are the category cutoffs for the control group for ease of visual comparison.

In Model 2, we added a variable testing for a conditional association between respondents who are politically conservative and viewed the *Female-Narr*-Video. The interaction term tests whether exposure to the video with the female narrator had different effects for people who identify as conservative than for those who do not identify as conservative. Political conservatives in the US have been found to have lower vaccine propensity, linked in part to patterns of trust in expertise (46). Specifically, conservatives often see science as needing to conform to beliefs about religious authority and common sense (44), which also provide similar bases for beliefs about hierarchical, traditional gender relations (47). Given the persistence of beliefs about gender and scientific ability (48-51) and findings that violating traditional gender beliefs can induce negative reactions, particularly among political conservatives (52), we test the possibility that the gender of the person delivering a message may affect how the message reaches political conservatives.

The addition of the politically conservative x *Female-Narr*-Video interaction variable showed that the main effect of the *Female-Narr*-Video for non-conservative respondents was a positive association consistent across levels of intent on the margin of statistical significance (coefficient = 0.374, *p* = 0.051). The magnitude of the coefficient is closer to the *Male-Narr*-Video (coefficient = 0.440, *p* = 0.005) than it was in Model 1 for any of the levels of intent, although the larger standard error for the *Female-Narr*-Video does suggest more heterogeneity remaining in the effect of the *Female-Narr*-Video. The interaction term shows a notable significant negative association for conservatives exposed to the *Female-Narr*-Video being less likely to express a vaccination intention greater than the “uncertain” response (coefficient = −0.971, *p* =.002) when compared to either conservatives in another of the experimental or control conditions or to others exposed to the *Female-Narr*-Video. In other words, among political conservatives, exposure to the video with a female narrator seemed to decrease the propensity of getting to a yes response (whether “probably yes” or “definitely yes”) for the vaccine when compared to political conservatives who viewed the *Male-Narr*-Video and even the control group.

Although the *Female-Narr*-Video had a backfire effect for conservatives, for other respondents the male- and female-narrated videos had relatively similar associations with increasing vaccination intention. These patterns are illustrated in Figure 2, which compares the predicted probabilities for a white, male participant, under the age of 55, with at least a BA, varying both the treatment and political ideology (comparing conservative to moderate). The figure shows that there is a negligible difference in predicted probabilities for a moderate or conservative exposed to the *Male-Narr*-Video of having either a “definitely yes” or “probably yes” response (83.0% and 81.4% respectively). The same comparison for the *Female-Narr*-Video shows a substantially wider gap (82.1% for moderates, 60.6% for conservatives). Notably, the predicted probability for a conservative exposed to the *Female-Narr-*Video to reply to the vaccination intention question with “definitely yes” or “probably yes” (60.6%) is also substantially lower than for a conservative in the control group that was not exposed to any information (73.7%). Tests for similar conditional effects for *Male-Narr*-Video on political conservatives showed no such conditional association (see SI Appendix). For example, adding an additional interaction term for conservative x *Male-Narr-*Video to the variables in Model 2 produced a coefficient with a near zero association (coefficient = −0.005, *p* = 0.987), while not changing any substantive patterns reported in Table 2.

**Figure 2.**
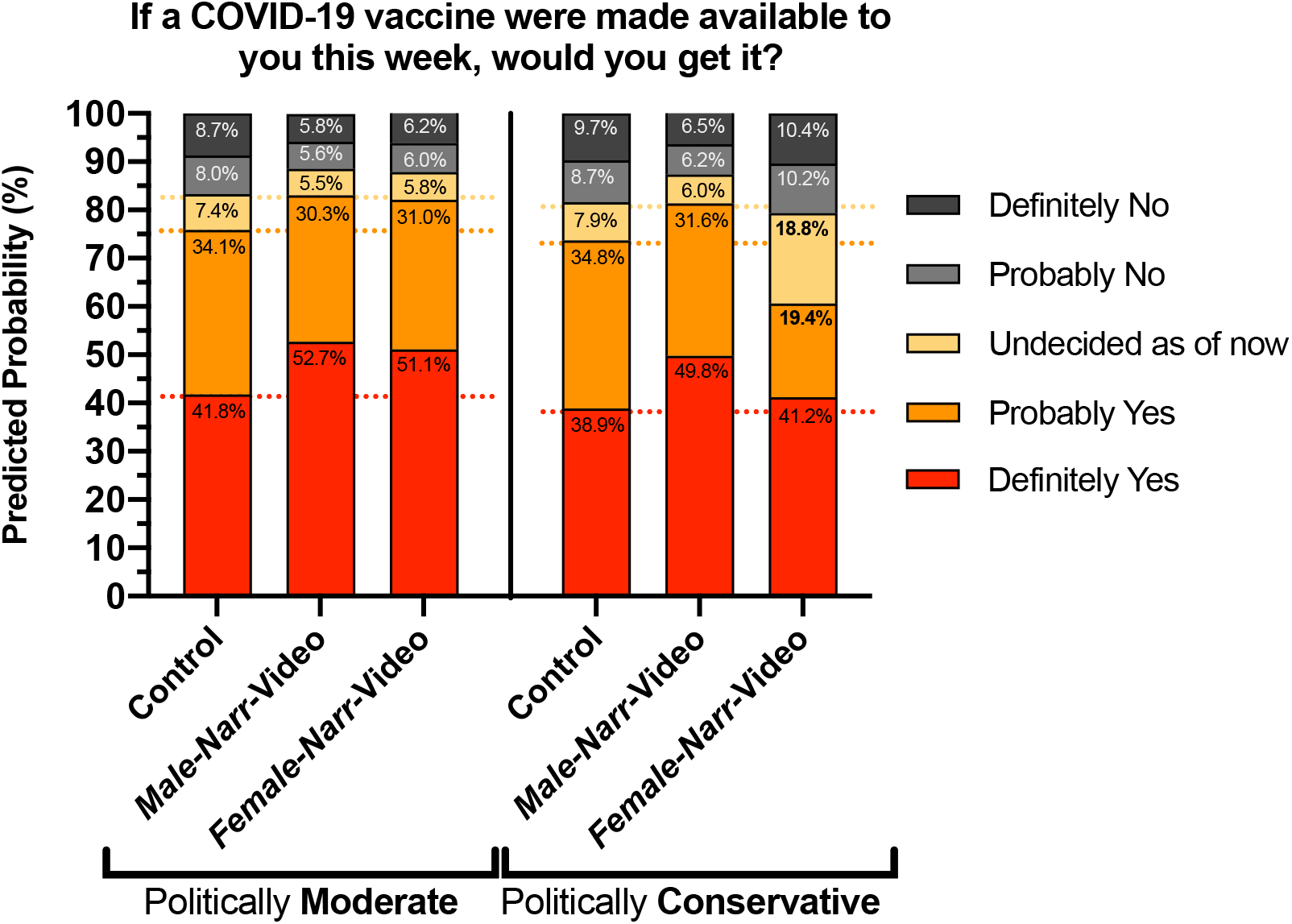
Predicted probabilities from Model 2 upon exposure to each of the four interventions for a participant who is white, male, under 55, with at least a B.A. degree, and comparing whether they identify as politically moderate or conservative. The horizontal dotted lines are the category cutoffs for the control group for ease of visual comparison.

### Perceptions of narrator quality

Despite the differences in effect on vaccination intention between the *Female-Narr*-Video and *Male-Narr*-Video, participants as a whole rated the quality of instruction in the two versions of the video equally highly. Contrary to previous research that has found that when quality of instruction is measured on a 10-point scale, gender gaps in favor of men are wider, particularly in male-dominated fields (53), there was an insignificant difference in the other direction among respondents included in the analysis. In a bivariate model, respondents who saw the *Female-Narr-*Video rated the quality of instruction 0.08 points higher, on average, on a 10-point scale than those who saw the *Male-Narr-*Video (p = 0.406, see Supplemental Table 2). Although conservatives rated the video about a quarter point lower than other respondents, there is no evidence of conditioning of conservatives’ rating based on which version of the video they saw— the nonsignificant coefficient for the interaction term between conservative and the *Female-Narr-* Video condition has a positive sign. The survey items on likelihood to share the video on social media, and the trustworthiness, comfort, and knowledge of the narrator (Supplemental Figures 2-3) likewise do not show evidence of explicit bias between the male and female-narrated versions of the video. Yet, the enhanced persuasive power of the *Male-Narr*-Video as seen in the increased intention to vaccinate, particularly for political conservatives, may be evidence of bias against women as sources of scientific authority, and is worthy of continued study.

## Conclusion

Widespread uptake of the COVID-19 vaccines worldwide is crucial for ending the COVID-19 pandemic. Previous work on vaccine hesitancy interventions have recommended that health organizations use social media channels such as YouTube to spread pro-vaccination messages, but caution that vaccine communications should first be tested (7, 19-21). Herein, we gathered empirical evidence on the effect of an 8-minute animated educational COVID-19 mRNA vaccine explainer video. We found that this video demonstrated a statistically significant association with increasing vaccination propensity, but that the messenger matters – the video with a male narrator had a robust association with increased vaccination intention, while the identical video with a female narrator had much more uneven associations, part of which could be accounted for by conservatives uniquely expressing uncertainty in vaccination intention after viewing that version of the video. The video studied herein only addresses the mRNA vaccines and does not explain many of the other vaccine questions and concerns people have. Future studies that follow up on the theoretical implications of these data may further examine the ways that gender beliefs influence how members of the public respond to communication about science and medicine, and help guide interventions promoting vaccine uptake.

## Data Availability

The dataset used for this analysis is available in the Supplemental information.

## Acknowledgments

The authors thank Dennis Cao for providing the voice of the male narrator. LSW is supported by a Cottrell Scholar Award from the Research Corporation for Science Advancement (ID No: 27499)

## Footnotes

† As the most direct measure of vaccine hesitancy is the vaccination intent survey item, the analyses in this paper focus on the responses to that question. Subsequent research may examine the other survey items in greater depth.

‡ This finding of no association between Latinx/Hispanic respondents and vaccine intent may be due to within-group variability that both associates with intent and decreases the likelihood of performing Mechanical Turk work tasks (39). Additionally, these models may account for some of the within-group variation, since 9% of in-sample respondents who identified at Latinx/Hispanic also identified as Black or Native American; previous research finds of visible markers of African and indigenous ancestry (40) can associate with health-related differences among Latinx/Hispanic people.

## Notes

### Competing Interest Statement

There are no competing interests to disclose. The authors have not received any payments or services from a third party in the past 36 months with entities that could be perceived to influence, or that give the appearance of potentially influencing, the submitted work. (LSW made the YouTube video as a part of a series of instructional videos made for educational purposes, and has not, and does not plan to, generate revenue from the videos in this study).

### Funding Statement

LSW is supported by a Cottrell Scholar Award from the Research Corporation for Science Advancement (funding started after the completion of this study ID No: 27499)

### Author Declarations

The study was approved by the Macalester College Institutional Review Board (Approval 022103)

