## Supplementary material for "A randomized controlled trial of a video intervention shows evidence of increasing COVID-19 vaccination intention": SI Appendix

### Supplementary Information for
A randomized controlled trial of a video intervention shows evidence of increasing COVID-19 vaccination intention.

Leah S. Witus* and Erik Larson*

* Corresponding authors: Leah S. Witus and Erik Larson

**This PDF file includes:**

- Supplementary text
- Tables S1 to S2
- Figures S1 to S3
- Code used in data analysis
- Full survey administered
- Legend for Dataset S1
- SI References

**Other supplementary materials for this manuscript include the following:**

- Dataset S1

**Supplementary text**

**Sensitivity analysis.** Additional analyses demonstrated that the findings discussed in the main text were robust to a variety of alternative specifications. We summarize these alternative specifications and describe what happens to the main coefficients of interest and any other substantial changes. While we include coefficients in this description, we emphasize that these coefficients should not be compared to the coefficients in the models in the main text. Rather, we include the coefficients below to allow for comparison of the relative difference of coefficients within particular models to the relative difference of the same coefficients in the models reported in the main text.

Across each of these sensitivity analyses, a single coefficient for *Male-Narr*-Video remains statistically significant and positively associated with increased vaccine intent relative to the control group across all levels of vaccine intent. Additionally, the pattern demonstrated in Model 2, showing a significant interaction term with a negative coefficient between *Female-Narr*-Video and politically conservative respondents is statistically significant for all analyses, although in two instances we departed from an autofit model (discussed below in further detail). The sensitivity analyses show that the Model 2 coefficient for the main effect of the *Female-Narr*-Video is most sensitive to the alternative specifications, often exhibiting greater difference from the *Male-Narr*-Video coefficient and moving further away from the cut-off from statistical significance. This pattern is consistent with the findings that the *Female-Narr*-Video associates with more varied response than the *Male-Narr*-Video and that some, but not all, of the heterogeneity relates to how political conservative identity conditions the response to *Female-Narr*-Video but not the *Male-Narr*-Video.

Partial proportional odds models can produce negative predicted probabilities for some cases (1). Eliminating four cases for which one predicted probability was negative did not change the results for Models 1 or 2.

As briefly summarized in the main text, we examined whether an interaction term between *Male-Narr*-Video and conservative respondents might illustrate any conditioning similar to what we found with the *Female-Narr*-Video. We tested this interaction term both by substituting it for the interaction term in Model 2 and by adding it as an additional variable along with the interaction term for *Female-Narr*-Video x conservative. When the *Male-Narr*-Video interaction term substituted for the *Female-Narr*-Video interaction term, the main effect coefficient for *Male-Narr*-Video remains statistically significant (coefficient = 0.394, $p$ = 0.041), while the main effect for conservative increases in magnitude relative to the *Male-Narr*-Video coefficient but is not statistically significant (coefficient = -0.253, $p$ = 0.131). Notably, the coefficient for the interaction term is far from statistically significant, but positive (coefficient = 0.119, $p$ = 0.678). When the *Male-Narr*-Video x conservative variable is added to the model along with the *Female-Narr*-Video x conservative variable, the relative magnitude of each of the main effect coefficients for these variables are essentially unchanged in relative magnitude compared to their relative magnitude in Model 2 as reported in the main text (*Male-Narr*-Video coefficient = 0.441, $p$ = 0.023; *Female-Narr*-Video coefficient = 0.375, $p$ = 0.054; conservative coefficient = -0.118, $p$ = 0.528). Similarly, the coefficient for the *Female-Narr*-Video x conservative interaction for the third level of the outcome (coefficient = -0.972, $p$ = 0.003) has a similar relative magnitude to the other coefficients in this model as it does in Model 2 in the main text. The coefficient for the *Male-Narr*-Video x conservative interaction term is nearly zero (coefficient = -0.005, $p$ = 0.987). This pattern increases confidence in the finding that there is something unique happening with the *Female-Narr*-Video, particularly with conservative respondents, rather than with the video treatment more generally.

Restricting analysis to a subset of respondents on the basis of race showed that the results were not the product of the heterogenous reference groups for the race and ethnicity variables in the main text models. Restricting analysis to only respondents who identified as white alone or black (whether alone or in combination with any other race or ethnicity) reduces the sample to 1,013. Doing so did not result in any coefficients from the models in the main text becoming statistically insignificant. There was some attenuation in the statistical significance for the *Male-Narr*-Video coefficient (e.g., in Model 2 the coefficient was 0.379, $p$ = 0.026). Notably, the main effect term for *Female-Narr*-Video had a larger difference from the *Male-Narr*-Video in Model 2 and was further from statistical significance (coefficient = 0.272, $p$ = 0.192).

In two alternate specifications, the autofit option results in a single coefficient for the interaction term between *Female-Narr*-Video and conservative respondents (negative, but not statistically significant), while producing different coefficients for each level of the outcome for the main effect of *Female-Narr*-Video. In these cases, the coefficient for the main effect *Female-Narr*-Video is statistically significant for responses greater than "probably yes" (the fourth and final level reported in the model). For both specifications, we also estimated a model imposing parallel lines (a single coefficient across all levels of vaccine intent) on the main effect variable. Since we are testing whether the earlier finding that some of the difference in response to the *Female-Narr*-Video associates with how conservative respondents may react with increased uncertainty, theory guides the imposition of the parallel line on the main effect variable (1). In both instances, when the parallel line is imposed on the main effect variable, the interaction term follows the pattern reported in Model 2 in the main text and AIC/BIC tests show support for the non-autofit model in which the main effect is subject to the parallel line imposition and the interaction term allowed to vary across levels of vaccine intent. Finally, predicted probability calculations show a similar pattern for both autofit and non-autofit models, particularly when comparing conservatives exposed to the two different video treatments.

Restricting the reference group for political ideology to only those who identified as moderate by dropping those who identified as something else results in the replicating the finding of a significant association for the *Male-Narr*-Video treatment variable and vaccine intent for both the autofit (coefficient = 0.472, $p$ = .003, all variables reported for Model 2) and parallel lines imposed on the *Female-Narr*-Video models (coefficient = 0.471, $p$ = 0.003). Under an autofit model, the *Female-Narr*-Video x conservative interaction has a single coefficient for all levels of the outcome (coefficient = -0.430, $p$ = 0.150), while the main effect coefficient for *Female-Narr*-Video has a statistically significant coefficient for a response above "probably yes" (coefficient = 0.472, p = 0.003). Maintaining the parallel line specification in Model 2 for the main effect for *Female-Narr*-Video (coefficient = 0.352, $p$ = 0.067) replicates the pattern in the model in the main text for the interaction term having a statistically significant association for the third level of the dependent variable (coefficient = -0.935, $p$ = .003). Comparing predicted probabilities for either a "probably yes" or "definitely yes" on vaccine intent shows that both these models reproduce the pattern reported for predicted probabilities for Model 2 in the main text. Using the same parameters as in the main text (a respondent under 55, white, male, B.A. or higher), we compare the predicted probabilities for conservatives exposed to both videos. Under the autofit model, a conservative exposed to the *Male-Narr*-Video has an 82.0% predicted probability of having a definitely or probably yes response, while one exposed to the *Female-Narr*-Video has a predicted probability of 64.6%. For the model with the parallel lines imposed, the comparison is 81.6% to 60.7%. Despite some difference in the magnitude of the difference, the substantive pattern and direction is similar. Finally, Akaike and Bayesian Information Criteria show a slight preference for the model for the imposition of parallel lines on the main effect (2955.824 for the non-autofit model compared to 2947.535 for the autofit model).

We find a similar pattern in broadening the sample to exclude only those who did not pass the attention check item, regardless of time spent with the video or text. Doing so increases the analytic sample to 1256 cases. Other than the pattern with the *Female-Narr*-Video interaction and main term in autofit models, this specification does not change the significance or direction of any coefficients, although the text intervention coefficient increases (coefficient = 0.219, p = 0.147) relative to the *Male-Narr*-Video coefficient (coefficient

= 0.440, p = 0.004). The pattern in the *Female-Narr*-Video and interaction term follows the pattern discussed in the previous paragraph: in the autofit model, the main effect variable has a significant positive association (coefficient = 0.399, *p* = 0.036) for the highest level outcome of the dependent variable. Following the pattern of Model 2 in the main text, in the model with parallel lines imposed on *Female-Narr*-Video, the interaction term has a significant negative association at the third level of the outcome (coefficient = -0.754, *p* = 0.011). Using the same method of comparing predicted probabilities as the previous paragraph for an under 55, white, male conservative with a B.A. or higher exposed to the *Male-Narr*-Video has an 81.2% predicted probability of having a definitely or probably yes response under the autofit model, compared to a predicted probability for exposure to the *Female-Narr*-Video has of 67.1%. The estimates from the model with parallel lines imposed are 80.9% and 64.0%, respectively. And, as above, the information criteria suggest the non-autofit model with parallel lines imposed is slightly better (3171.610 for the parallel lines model, compared to 3171.909 for the autofit model).

Analyses that restricted the sample based on level of education suggested that the associations reported in Models 1 and 2 in the main text may reflect somewhat distinct processes based on respondents' level of education. When Model 1 was restricted only to respondents with an education level lower than a B.A., the *Male-Narr*-Video group (coefficient = 0.744, p = 0.004) and *Female-Narr*-Video group (coefficient = 0.078, p = 0.770) both had coefficients with symmetrical associations across the different levels of vaccine intent. More notably, the magnitude of difference between the two coefficients is substantially increased. In Model 2, the coefficients for both video treatments, political conservatives, and the interaction between conservative and *Female-Narr*-Video all had symmetrical associations with vaccine intent. While the coefficients for both political conservatives (coefficient = -0.382 p = 0.153) and the interaction term (coefficient = -0.356, p = 0.491) were negative, neither is statistically significant, nor is the coefficient for *Female-Narr*-Video (coefficient = 0.180, p = 0.556). It should be noted, however, that the statistical power for this model is substantially attenuated, since it includes only 395 cases.

When restricted only to respondents with an education level of a B.A. or higher, neither video treatment coefficient is statistically significant in Model 1 and the two coefficients are of a similar magnitude (*Male-Narr*-Video coefficient = 0.244, p = 0.216; *Female-Narr*-Video coefficient = 0.249, p = 0.207). In Model 2, however, the coefficient for interaction term for *Female-Narr*-Video and conservative is of a notable magnitude for a response higher than "undecided as of now" (coefficient = –1.150, p = 0.003), reflecting the pattern in the main text model. Additionally, the interaction term's coefficient for at the second level (higher than "probably not") is negative and modestly above the cut off for statistical significance (coefficient = –0.788, p = 0.064). Including these interaction terms in the model affects the coefficient for the main term for *Female-Narr*-Video (for respondents other than conservatives). It is increased in magnitude and statistically significant (coefficient = 0.517, p = 0.038). The coefficient for conservative is essentially zero (coefficient = 0.018, p = 0.930). As above, this model has reduced statistical power due in part to the reduced sample size and in part due to the lower level of variation in vaccine intent among those with a B.A. or higher in the sample.

The findings from these sample restrictions on the basis of education may, therefore, reflect a specification pattern, finding a greater difference between the two video treatments across respondents with lower levels of education and, at higher levels of education, particularly strong evidence of the conditioning of the effect of the gender of the narrator based on political ideology. In other words, the impact of the gender of the video narrator on vaccination intention might reflect a pattern of the *Female-Narr*-Video being less persuasive to those with a lower level of education and a pattern of more educated conservatives reacting negatively to the *Female-Narr*-Video.

**Supplemental Table 1:** Frequencies for variables included in analytic sample (n=1184)

| | |
|---|---|
| *Dependent Variable: If a COVID-19 vaccine were made available to you this week, would you get it?* | |
| Definitely no | 106 (9.0%) |
| Probably no | 96 (8.1%) |
| Undecided as of now | 97 (8.2%) |
| Probably yes | 265 (22.4%) |
| Definitely yes | 620 (52.4%) |
| *Treatment Condition* | |
| Video Male Narrator | 270 (22.8%) |
| Video Female Narrator | 254 (21.5%) |
| Text/Blog Post | 276 (22.3%) |
| *Demographic Controls* | |
| Female | 520 (43.9%) |
| 55 or older | 158 (13.3%) |
| Black/African-American | 117 (9.9%) |
| American Indian/Alaska Native | 22 (1.9%) |
| Politically conservative | 376 (31.8%) |
| Politically liberal | 578 (48.8%) |
| Highest education: B.A. or higher | 789 (66.6%) |

**Supplemental Table 2:** Linear regression model of the survey item rating the quality instruction on a 10-point scale considering political identification (standard errors in parentheses). The data in the table includes only cases from the analytic sample that were exposed to one of the two video conditions.

| | Model 1 | Model 2 | Model 3 |
|---|---|---|---|
| *Female-Narr*-Video | 0.101 | 0.091 | 0.083 |
| (compared to *Male-Narr*-Video) | (0.107) | (0.107) | (0.130) |
| Conservative | | -0.210 | -0.221 |
| (compared to all others) | | (0.114) | (0.156) |
| | | *p*=.067 | |
| *Female-Narr*-Video x | | | 0.024 |
| Conservative | | | (0.229) |
| Constant | 9.090 | 9.162 | 9.167 |

N=517

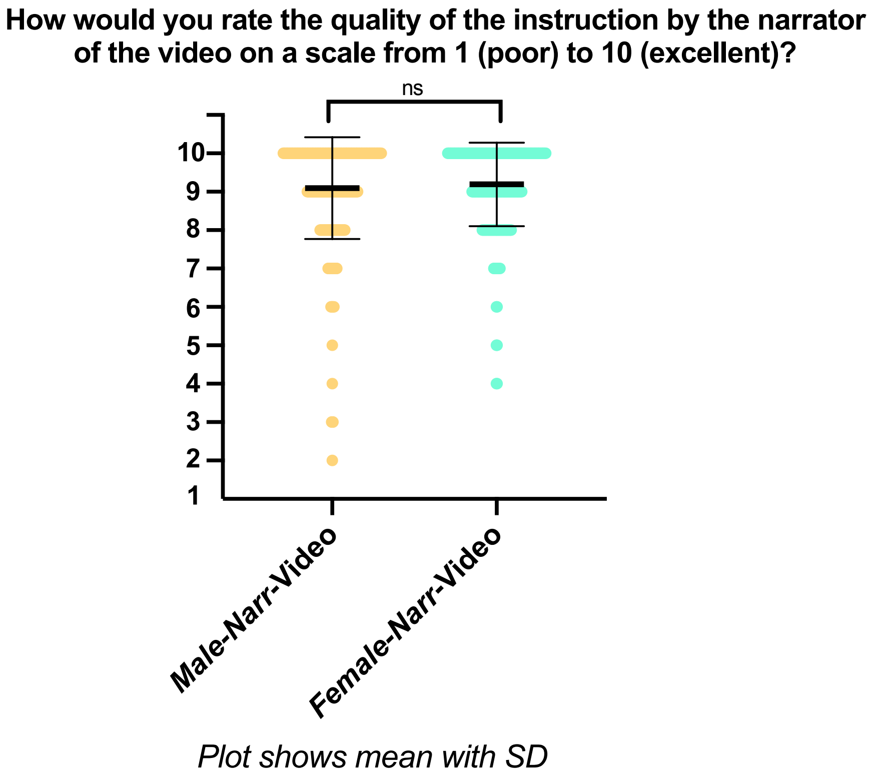

**Supplemental Figure 1:** Survey responses to the question "How would you rate the quality of the instruction by the narrator of the video on a scale from 1 (poor) to 10 (excellent)?". Significance indicated based on a Mann Whitney test in GraphPad Prism 9.

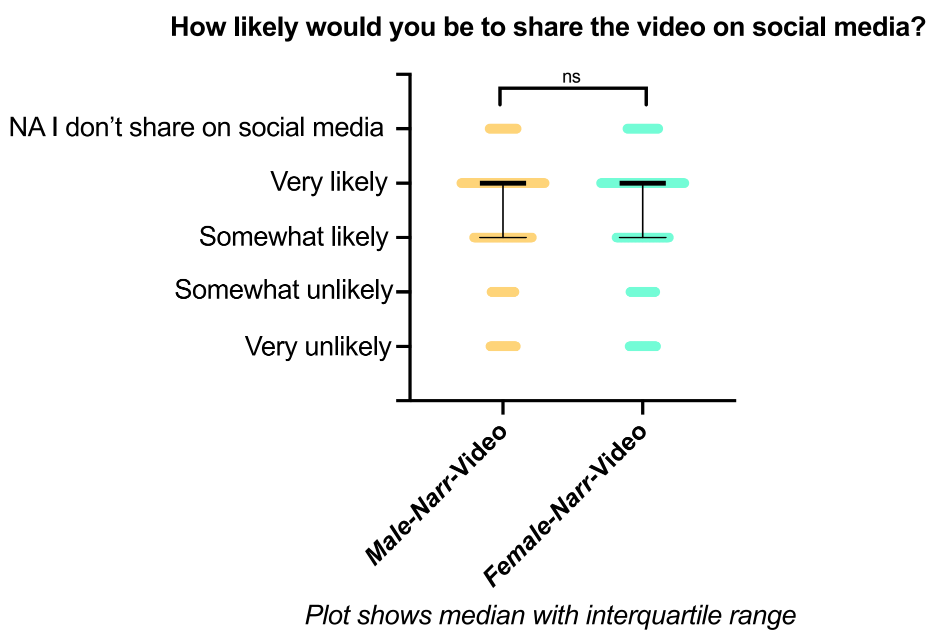

**Supplemental Figure 2:** Survey responses to the question "How likely would you be to share the video on social media?". Significance indicated based on a Mann Whitney test in GraphPad Prism 9.

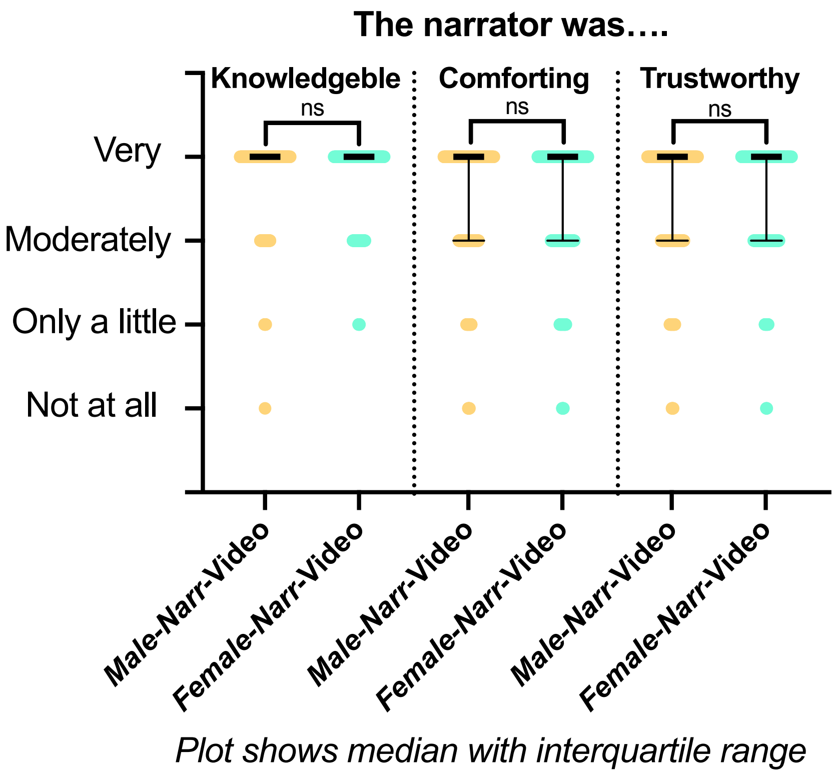

**Supplemental Figure 3:** Survey responses to the questions regarding narrator knowledge, trustworthiness, and comfort. Significance indicated based on a Mann Whitney test in GraphPad Prism 9.

**Code used for data analysis**

We imported the data from Qualtrics to SPSS for initial processing and defining all variables. We then exported the SPSS file into Stata for analysis. We, therefore, include SPSS codes for the data processing and Stata code for the data analysis.

```
**SPSS Data Preparation

**Creating variable for groups

RECODE Q37_Page_Submit (MISSING=99) (1 thru Highest=1) (ELSE=99) INTO Group.
VARIABLE LABELS  Group 'Which group'.
EXECUTE.
DO IF (Q36_Page_Submit>0).
RECODE Group (99=2).
END IF.
EXECUTE.
DO IF (Q38_Page_Submit>0).
RECODE Group (99=3).
END IF.
EXECUTE.
DO IF (Q37_Page_Submit>0).
RECODE Group (99=4).
END IF.
EXECUTE.

**Create dummy variables for treatment conditions

RECODE Group (1=1) (MISSING=SYSMIS) (ELSE=0) INTO FNV.
VARIABLE LABELS  FNV 'Video group female narrator'.
EXECUTE.
RECODE Group (2=1) (MISSING=SYSMIS) (ELSE=0) INTO MNV.
VARIABLE LABELS  MNV 'Video group male narrator'.
EXECUTE.
RECODE Group (3=1) (MISSING=SYSMIS) (ELSE=0) INTO text.
VARIABLE LABELS  text 'Text group'.
EXECUTE.

**Create dummy variable for respondents identifying as female

RECODE Q27 (2=1) (MISSING=SYSMIS) (ELSE=0) INTO Female.
VARIABLE LABELS  Female 'Respondent identified as female'.
EXECUTE.

** Create variable for outcome excluding those already vaccinated

RECODE Q8 (6=SYSMIS) (ELSE=Copy) INTO Q8R.
VARIABLE LABELS  Q8R 'If vaccine offered this week excluding those already vaccinated'.
EXECUTE.
```

```
**Age recode to create 55 or older dummy

RECODE Q29 (MISSING=SYSMIS) (6 thru 9=1) (ELSE=0) INTO A55Plus.
VARIABLE LABELS  A55Plus 'Age is 55 or older'.

**Pol ideol dummies

RECODE Q35 (MISSING=SYSMIS) (1=1) (2=1) (3=1) (ELSE=0) INTO PolCon.
VARIABLE LABELS  PolCon 'Describes self as politically conservative'.
EXECUTE.
RECODE Q35 (MISSING=SYSMIS) (5=1) (6=1) (7=1) (ELSE=0) INTO PolLib.
VARIABLE LABELS  PolLib 'Describes self as politically liberal'.
EXECUTE.

**Educ level recode--binary BA+/less than BA

RECODE Q37 (MISSING=SYSMIS) (6=1) (7=1) (ELSE=0) INTO BAPlus.
VARIABLE LABELS  BAPlus 'Highest level of ed BA or beyond'.
EXECUTE.

**Race dummies--not forcing mutual exclusivity other than for white only

RECODE Q39_2 Q39_6 Q39_3 Q39_4 Q39_5 Q39_7 (MISSING=0) (1=1) INTO Black Latino Indig Asian
   PacIsle OthRace.
VARIABLE LABELS  Black 'Identiifes as black alone or with other' /Latino 'Indentifies Latino or '+
   'Hispanic' /Indig 'Identifies Native American' /Asian 'Identifies Asian' /PacIsle
   'Identifies Hawaiian or Pacific Islander' /OthRace 'Identifies some other race'.
EXECUTE.

**Create interaction variables

COMPUTE FNVCon=FNV*PolCon.
VARIABLE LABELS  FNVCon 'FNV by Conservative Interaction'.
EXECUTE.

COMPUTE MNVCon=MNV*PolCon.
VARIABLE LABELS  MNVCon 'MNV by Conservative Interaction'.
EXECUTE.

**Primary exclusion inclusion criterria--if less than 7 min (start of conclusion) and 2 min 5 sec on reading
task (similar percentage exclsion) OR if attention check incorrect
** Include if SampleA = 0

RECODE Q36_Page_Submit (MISSING=0) (Lowest thru 420=1) (ELSE=0) INTO LT7.
VARIABLE LABELS  LT7 'Clicked from video in less than 7 min or blog in 125 sec'.
EXECUTE.
```

```
DO IF (Q37_Page_Submit<420).
RECODE LT7 (0=1).
END IF.
EXECUTE.

DO IF (Q38_Page_Submit<125).
RECODE LT7 (0=1).
END IF.
EXECUTE.

RECODE LT7 (ELSE=COPY) INTO SampleA.
VARIABLE LABELS SampleA 'Exclude if short time or attention check incorrect'.
EXECUTE.

DO IF (Q19 = 1).
RECODE SampleA (0=1).
END IF.
EXECUTE.

DO IF (Q22 = 1).
RECODE SampleA (0=1).
END IF.
EXECUTE.

** Secondary: exclude only if attentnion question is wrong
** Include if SampleB=0

RECODE Q22 (2=0) (1=1) (MISSING=0) INTO SampleB.
VARIABLE LABELS  SampleB 'If attention question was incorrect exclude'.
EXECUTE.

DO IF (Q19 = 1).
RECODE SampleB (0=1).
END IF.
EXECUTE.
```

Stata Code for analysis

```
* set missing value for dependent variable

mvdecode Q8R, mv(6)

* Main model

nestreg: gologit2 Q8R (MNV FNV text BAPlus Black Indig A55Plus Female PolCon PolLib) (FNVCon)
if(SampleA==0), autofit

* test robustness dropping any case with any negative predicted probability

predict m2a if e(sample)==1, outcome(1)
predict m2b if e(sample)==1, outcome(2)
predict m2c if e(sample)==1, outcome(3)
predict m2d if e(sample)==1, outcome(4)

*only m2d/outcome 4 had any negative values—test to see if excluding those cases causes substantive
change

nestreg: gologit2 Q8R (MNV FNV text BAPlus Black Indig A55Plus Female PolLib PolCon) (FNVCon)
if(SampleA==0 & m2d>=0), autofit

* test MNV x Conservative interaction: first substitute for FNVCon, then both together

gologit2 Q8R MNV FNV text BAPlus Black Indig A55Plus Female PolCon PolLib MNVCon
if(SampleA==0), autofit

gologit2 Q8R MNV FNV text BAPlus Black Indig A55Plus Female PolCon PolLib MNVCon FNVCon
if(SampleA==0), autofit

* check if rating of quality instruction varies by FNV

nestreg: regress Q26 (FNV) (PolCon) (FNVCon) if !missing(m2a)

* test by drop to white alone or black alone

nestreg: gologit2 Q8R (MNV FNV text BAPlus Black Indig A55Plus Female PolLib PolCon) (FNVCon) if
(SampleA==0 & Latino!=1 & Indig!=1 & Asian!=1 & PacIsle!=1 & OthRace!=1), autofit

* test alternate specification of pol ideology: due to autofit pattern, run non-nested and compare

gologit2 Q8R MNV FNV text BAPlus Black Indig A55Plus Female PolCon PolLib if(SampleA==0 &
Q35<8), autofit

gologit2 Q8R MNV FNV FNVCon text BAPlus Black Indig A55Plus Female PolCon PolLib if(SampleA==0
& Q35<8), autofit
```

```
fitstat, save

gologit2 Q8R MNV FNV FNVCon text BAPlus Black Indig A55Plus Female PolCon PolLib if(SampleA==0
& Q35<8), pl (MNV FNV text Black Indig Female PolCon PolLib)

fitstat, diff

* test only BA+ and only <BA

nestreg: gologit2 Q8R (MNV FNV text Black Indig A55Plus Female PolCon PolLib) (FNVCon)
if(SampleA==0 & BAPlus==0), autofit

nestreg: gologit2 Q8R (MNV FNV text Black Indig A55Plus Female PolCon PolLib) (FNVCon)
if(SampleA==0 & BAPlus==1), autofit

* exclude only on basis of attention check

gologit2 Q8R MNV FNV text BAPlus Black Indig A55Plus Female PolCon PolLib if(SampleB==0), autofit

gologit2 Q8R MNV FNV FNVCon text BAPlus Black Indig A55Plus Female PolCon PolLib
if(SampleB==0), autofit

fitstat, save

gologit2 Q8R MNV FNV FNVCon text BAPlus Black Indig A55Plus Female PolCon PolLib
if(SampleB==0), pl (MNV FNV text Black Indig Female PolCon PolLib)

fitstat, diff
```

**Full Survey**

#### Survey Flow

EmbeddedData
Random ID = ${rand://int/10000:99999}

Block: Introductory text (1 Question)

BlockRandomizer: 1 - Evenly Present Elements

Group: Male video

Standard: Male video (2 Questions)
Block: COVID vaccine questions (6 Questions)
Block: Video questions (6 Questions)
Block: Demographics (8 Questions)

Group: Female video

Standard: Female video (2 Questions)
Standard: COVID vaccine questions (6 Questions)
Standard: Video questions (6 Questions)
Standard: Demographics (8 Questions)

Group: Text

Standard: Blog post (text) (2 Questions)
Block: COVID vaccine questions (6 Questions)
Standard: Text questions (6 Questions)
Block: Demographics (8 Questions)

Group: Control

Block: COVID vaccine questions (6 Questions)
Block: Demographics (8 Questions)
Standard: Dont need to watch (1 Question)

EmbeddedData
Random ID = ${e://Field/Random%20ID}

Block: completion code (1 Question)

Page Break

Start of Block: Introductory text

Q1 *What*: We are conducting this survey to examine how different methods of communication affect how people understand the COVID19 mRNA vaccines. We will share information with you in one randomly assigned format and ask you a series of short questions. The entire process will take less than 20 minutes to complete.

*Who*: This survey is being conducted by Macalester College Professors Leah Witus and Erik Larson. If you wish to contact us, you may do so at.

*Data Use and Confidentiality*: We are not collecting any names or email addresses as part of this survey and will not know your identity. Your responses will be only be seen by Leah Witus and Erik Larson, who will analyze the results. Aggregate results may be included in a publication in a scholarly journal. Data will be stored securely and managed by Leah Witus and Erik Larson.

*Participants*: This survey is being administered via Amazon Mechanical Turk. Responding to this survey is voluntary and even if you decide to participate, you may decide to withdraw at any point. You will receive payment via Amazon Mechanical Turk by entering the code you receive after the completion of this survey.

End of Block: Introductory text

---

Start of Block: Male video

Q5 The video below was created by a college biochemistry professor.
Please watch the entirety of the following video.
https://youtu.be/Fv5bs4SPiYE

---

Q36 Timing
First Click (1)
Last Click (2)
Page Submit (3)
Click Count (4)

End of Block: Male video

---

Start of Block: COVID vaccine questions

Q8 If a COVID-19 vaccine were made available to you this week, would you get it?

- Definitely no (1)
- Probably no (2)
- Undecided as of now (3)
- Probably yes (4)
- Definitely yes (5)
- I already got one or more dose of a COVID19 vaccine (6)

---

Q10 How well do you understand the following?

| | Not at all (1) | Only a little (2) | Moderately (3) | Well (4) | Very well (5) |
|---|---|---|---|---|---|
| I understand how vaccination rates affect herd immunity (4) | ○ | ○ | ○ | ○ | ○ |
| I understand what a virus is (3) | ○ | ○ | ○ | ○ | ○ |
| I understand how vaccines work (1) | ○ | ○ | ○ | ○ | ○ |
| I understand what mRNA is (2) | ○ | ○ | ○ | ○ | ○ |

---

Q12 Please answer the following questions about the safety and efficacy of the COVID-19 mRNA vaccines

| | Not at all (1) | Only a little (2) | Moderately (3) | Very (4) | Completely (5) |
|---|---|---|---|---|---|
| To what extent are the COVID-19 mRNA vaccines **safe**? (5) | ○ | ○ | ○ | ○ | ○ |
| To what extent are the COVID-19 mRNA vaccines **effective**? (15) | ○ | ○ | ○ | ○ | ○ |

Q14 Please answer the following questions

| | Not at all (1) | Only a little (2) | Moderately (3) | A great deal (4) |
|---|---|---|---|---|
| To what extent is getting vaccinated important for the health of others in my community? (7) | ○ | ○ | ○ | ○ |
| To what extent are you concerned about other people not getting vaccinated? (16) | ○ | ○ | ○ | ○ |
| To what extent are you worried about potential side effects of a COVID-19 vaccine? (20) | ○ | ○ | ○ | ○ |

Q16 How much do you trust the following groups to act in the best interest of the public?

| | Not at all (1) | Only a little (2) | Moderately (3) | A great deal (4) |
|---|---|---|---|---|
| I trust the **medical community** to act in the best interests of the public (1) | ○ | ○ | ○ | ○ |
| I trust the **scientific community** to act in the best interests of the public (2) | ○ | ○ | ○ | ○ |
| I trust the **government regulators who approve vaccines** to act in the best interests of the public (3) | ○ | ○ | ○ | ○ |
| I trust the **companies that develop vaccines** to act in the best interests of the public (4) | ○ | ○ | ○ | ○ |

Q18 Rate the degree to which you find the following statements to be accurate or inaccurate

| | Inaccurate (1) | Somewhat inaccurate (2) | Neutral (3) | Somewhat accurate (4) | Accurate (5) |
|---|---|---|---|---|---|
| mRNA is an unproven vaccine technology (8) | o | o | o | o | o |
| New vaccines carry more risks than older vaccines (9) | o | o | o | o | o |
| Natural immunity is better than vaccine-acquired immunity (11) | o | o | o | o | o |
| Vaccines contain harmful ingredients (12) | o | o | o | o | o |

End of Block: COVID vaccine questions

Start of Block: Video questions

Q20 The following questions are on the video that you watched.

Q22 True or false? The video included information on the AstraZeneca adenovirus vaccine

▼ True (1) ... False (2)

Q24 How likely would you be to share the video on social media?

▼ Very unlikely (1) ... Not applicable (I don't share on social media) (5)

Q26 The creator of the video also narrated it.

How would you rate the **quality of the instruction** by the video creator on a scale from 1 (poor) to 10 (excellent)?

- ○ Poor 1 (1)
- ○ 2 (2)
- ○ 3 (3)
- ○ 4 (4)
- ○ 5 (5)
- ○ 6 (6)
- ○ 7 (7)
- ○ 8 (8)
- ○ 9 (9)
- ○ Excellent 10 (10)

Q28 Please comment on the characteristics of the narrator

| | Not at all (1) | Only a little (2) | Moderately (3) | Very (4) |
|---|---|---|---|---|
| The narrator was **trustworthy** (1) | ○ | ○ | ○ | ○ |
| The narrator was **comforting** (2) | ○ | ○ | ○ | ○ |
| The narrator was **knowledgeable** (3) | ○ | ○ | ○ | ○ |

Q30 Please rate the video on the following criteria

| | Not at all (1) | Only a little (2) | Moderately (3) | Very (4) |
|---|---|---|---|---|
| The video was **enjoyable** (1) | ○ | ○ | ○ | ○ |
| The video was **informative** (2) | ○ | ○ | ○ | ○ |
| The information presented in the video was **clear** (3) | ○ | ○ | ○ | ○ |

**End of Block: Video questions**

**Start of Block: Demographics**

Q25 Demographic questions

Q27 What is your gender?

▼ Male (1) ... Prefer not to say (4)

Q29 What is your age?

▼ Under 18 (1) ... 85 or older (9)

Q31 What is your annual household income level?

▼ Under $25,000 (1) ... Over $100,000 (5)

Q33 Do you have children?

▼ Yes - and at least one is under 18 years old (1) ... No (3)

Q35 How do you describe yourself politically?

▼ Extremely conservative (1) ... Other (8)

Q37 What is your highest level of education?

▼ Lower than high school (1) ... Other (8)

Q39 What is your race? Check all that apply

- ☐ White (1)
- ☐ Black or African American (2)
- ☐ American Indian or Alaska Native (3)
- ☐ Asian (4)
- ☐ Native Hawaiian or Pacific Islander (5)
- ☐ Latino/Hispanic (6)
- ☐ Other (7)

**End of Block: Demographics**

---

**Start of Block: Female video**

Q4 The video below was created by a college biochemistry professor.
Please watch the entirety of the following video.
https://youtu.be/j3hTeDyvgPs

Q37 Timing
First Click (1)
Last Click (2)
Page Submit (3)
Click Count (4)

**End of Block: Female video**

---

**Start of Block: Blog post (text)**

Q35 ***The blog post below was created by a college biochemistry professor.***

***Please read the entirety of the following text.***

To understand the COVID-19 vaccines and vaccination in general, it's helpful to review some core principles of biology.

1. Our bodies are made up of billions of cells. Inside cells, proteins are the biomolecules that are carrying out all of the different tasks that cells need to do to stay alive.
2. A cell knows which proteins to make from its DNA. Your DNA is stored in the nucleus of the cell.
3. When the cell needs to make a protein, it copies part of the instructions in the DNA into messenger RNA, mRNA, which is used by the cell's protein production machines to make the right proteins.
4. mRNAs are temporary instructions, they break down after use. Think of it like a grocery list written on a piece of paper, telling you what you need at the store – it doesn't stick around long after it's been used.

Okay, so that's how normal cells work. So what is a virus?

1. It's a small particle that's missing most of the components of a living cell.
2. Most viruses are just DNA or RNA inside a coating of proteins or fats. The viral RNA stores the instructions to make proteins, but the virus can't make them on its own. So that's why viruses need us to replicate.
3. Viruses make their way into healthy human cells and takeover. They send the instructions from their RNA or DNA to our protein production machines, and all of a sudden, our cells are making the virus's proteins, which assemble into new virus particles, which go out to infect more cells. As the viruses takeover our cells, it makes us feel ill.

But our immune system has ways to fight back against viruses.

1. Once the immune system has detected a viral invader, it produces proteins called antibodies that can recognize and neutralize the virus to prevent future attack.
2. In the case of COVID-19, the antibodies recognize the spike protein on the surface of the virus.
3. So that means for most viruses, if you've gotten them once you will have immunity going forward. If you're exposed to the virus again in the future, you won't get sick because you already have antibodies to prevent the virus from entering your cells and replicating.

That's a simplified picture of how the immune system normally works. How does this relate to vaccines?

1. Well throughout history people have wondered if there would be a way to boost your immune system to give it those antibodies (that recognize a virus and stop it from

replicating) without having to actually get sick from the virus first. Some viruses are unpleasant but you recover quickly, like cold viruses.

2. But some viruses like smallpox, measles, and COVID-19 can be deadly. So it's not worth risking getting sick.

*There is a way to train your immune system to make the right virus-fighting antibodies without getting sick first*. And that's what a vaccine is.

Vaccines boosts your immune system so that if you are exposed to a virus you already have the right antibodies ready to stop the virus from replicating and prevent you from getting sick.

1. Vaccines have been used for over 100 years. Back in the day, the vaccines were weakened versions of the actual virus.
2. This initial form of vaccination was admittedly tricky, because the weakened virus given as a vaccine had to be close enough to the actual virus so that the antibodies made by the immune system would work to prevent infection if the person was exposed to the real virus.
3. But the weakened virus vaccine had to be different enough from the real virus so that the vaccine itself would not infect people and make them sick.

That was vaccine technology 100 years ago. But today, thanks to decades of biomedical research, we have more options for making safe vaccines.

So how does the COVID-19 vaccine work? The first two vaccines developed for COVID-19, from the companies Pfizer and Moderna, are mRNA vaccines instead of weakened virus vaccines.

This is part of the reason they were able to be developed so quickly: instead of needing time to find the delicate balance between being close enough to the actual virus to produce the right antibodies, but weakened enough to not cause infection, mRNA vaccines are clever, because they're not viruses at all. They're just mRNA with the instructions for one part of the virus. And do they work?

When our cells encounter the vaccine mRNA, they follow the mRNA instructions and a protein that's a piece of the virus, the spike protein, is made. The good news is that this piece of the virus is enough for our immune system to produce antibodies to recognize the virus.

1. But the spike protein produced by the mRNA is not nearly enough to be infectious on its own. So there's no risk of infection from the vaccine.
2. The Pfizer and Moderna vaccines have been found to be highly effective in preventing illness from COVID-19. And if you remember, mRNA doesn't stick around long after it's been used. So it's very unlikely to have any long term side effects. mRNA vaccines are an amazing new technology, but it's still good practice to be cautious. Vaccine candidates are tested in 10s of thousands of clinical trial volunteers.
3. The COVID-19 mRNA vaccines were found to be highly effective (95% effective), with common minor side effects, such as soreness and tiredness, that are to be expected, as

your immune system is working hard to produce antibodies after you get the vaccine. A very small number of people have had other side effects.

It may be reassuring to know that the safety standards for vaccines are even higher than for other medicines.

Getting a vaccine is truly like gaining a superpower – it trains your own immune system to fight viruses and protects you from getting sick.

One more note about vaccines. You may hear talk of vaccination rates. Some people call this herd immunity.

1. The idea is that there are some people in any society that cannot get a vaccine for a particular virus. This includes small babies, and people with immune disorders.
2. So how can we protect these people that are not able to get a vaccine? Well if the vast majority of people in a population do get vaccinated, the virus will not be able to spread, because it can't travel from person to person, since most people have antibodies. This protects the people that can't get vaccinated.
3. That's why, even if you are not particularly worried about getting sick from a certain virus, you can still do a societal good deed if you get vaccinated.

So to sum it up, mRNA vaccines are a new type of safe, and highly effective vaccine, getting vaccinated for COVID-19, and other diseases, is a way to train and boost your body's natural immune system and it could save another person's life. Not bad.

Q38 Timing
First Click (1)
Last Click (2)
Page Submit (3)
Click Count (4)

**End of Block: Blog post (text)**

**Start of Block: Text questions**

Q18 The following questions are on the blog posted that you read.

Q19 True or false? The blog post included information on the AstraZeneca adenovirus vaccine

▼ True (1) ... False (2)

Q20 How likely would you be to share the blog post on social media?

▼ Very unlikely (1) ... Not applicable (I don't share on social media) (5)

Q21 How would you rate the **quality of the instruction** by the author of the blog post on a scale from 1 (poor) to 10 (excellent)?

- Poor 1 (1)
- 2 (2)
- 3 (3)
- 4 (4)
- 5 (5)
- 6 (6)
- 7 (7)
- 8 (8)
- 9 (9)
- Excellent 10 (10)

Q22 Please comment on the characteristics of the author of the blog post

| | Not at all (1) | Only a little (2) | Moderately (3) | Very (4) |
|---|---|---|---|---|
| The author was **trustworthy** (1) | ○ | ○ | ○ | ○ |
| The author was **comforting** (2) | ○ | ○ | ○ | ○ |
| The author was **knowledgeable** (3) | ○ | ○ | ○ | ○ |

Q23 Please rate the text of the blog post on the following criteria

| | Not at all (1) | Only a little (2) | Moderately (3) | Very (4) |
|---|---|---|---|---|
| The text was **enjoyable** (1) | ○ | ○ | ○ | ○ |
| The text was **informative** (2) | ○ | ○ | ○ | ○ |
| The information presented in the text was **clear** (3) | ○ | ○ | ○ | ○ |

**End of Block: Text questions**

**Start of Block: Dont need to watch**

Q40 Thank you for answering these questions. We have enough respondents in the group assigned to watch the scientific communication and do not need you to engage in that part of the survey. Please continue to submit the questions you have answered already to receive payment through Mechanical Turk.

**End of Block: Dont need to watch**

**Start of Block: completion code**

Q33
Thank you for your completion of this survey. Below is your MTurk Completion Code. You will need to enter this code in the Amazon Mechanical Turk interface to indicate your completion of this survey. After you record this code, be sure to click the next arrow below to submit your survey answers.

Copy this code to paste into MTurk:

${e://Field/Random%20ID}

Once you have copied this code, be sure to click the next arrow to submit your survey.

**End of Block: completion code**

---

**Legend for dataset S1**
The dataset .csv file includes the raw data from the survey items and variables defined for analysis in this paper.
